# Wastewater surveillance pilot at US military installations: a cost model analysis

**DOI:** 10.1101/2023.11.14.23298310

**Authors:** Jaleal Sanjak, Erin M McAuley, Richard Pinkham, Jacob Tarnowski, Nicole Miko, Bridgette Rasmussen, Christian Manalo, Michael Goodson, Blake Stamps, Bryan D Necciai, Shanmuga Sozhamannan, Ezekiel J Maier

## Abstract

**Background:** The coronavirus disease 2019 (COVID-19) pandemic highlighted the need for pathogen surveillance systems to augment both early warning and outbreak monitoring/control efforts. Wastewater samples provide a rapid and accurate source of environmental surveillance data to complement direct patient sampling. Due to its global presence and critical missions, the US military is a leader in global pandemic preparedness efforts. Clinical testing for COVID-19 on US Air Force (USAF) bases (AFBs) was effective, but costly with respect to direct costs and indirect costs of lost time. To remain operating at peak capacity such bases sought a more passive surveillance option and piloted wastewater surveillance (WWS) at 17 AFBs to demonstrate feasibility, safety, and utility from May 2021 to January 2022.

**Objective:** Here we model the costs of a wastewater program for pathogens of pandemic potential within the specific context of US military installations using assumptions based on the results of the USAF and Joint Program Executive Office for Chemical, Biological, Radiological and Nuclear Defense (JPEO-CBRND) pilot program. The objective was to determine the cost of deploying WWS to all AFBs, relative to clinical swab testing surveillance regimes.

**Methods:** A simple WWS cost projection model was built based on subject matter expert input and actual costs incurred during a WWS pilot program at USAF AFBs. Several SARS-CoV-2 circulation scenarios were considered and costs of both WWS and clinical swab testing were projected. Break even analysis was conducted to determine how reduction in swab testing could open up space to enable WWS to occur in complement.

**Results:** Our model confirms that wastewater surveillance is complementary and highly cost-effective when compared to existing alternative forms of biosurveillance. We find that the cost of WWS was between $10.5 - $18.5 million less expensive annually in direct costs as compared to clinical swab testing surveillance. When indirect cost of lost work is incorporated, including assumed lost work required to go obtain a clinical swab test, we estimate that over two thirds of clinical swab testing could be maintained with no additional costs upon implementation of WWS.

**Conclusions:** Our results support adoption of wastewater surveillance across US military installations as part of a more comprehensive and early warning system that will enable adaptive monitoring during disease outbreaks.

## Introduction

Many human pathogens are shed into bodily fluids during active infection and make their way into the sewage system along several routes. Therefore, wastewater sample collection is a viable approach to monitor for prevalence of pathogens [1], including those of pandemic potential and biodefense/biosecurity relevance. Early in the COVID-19 pandemic researchers identified that severe acute respiratory syndrome coronavirus 2 (SARS-CoV-2) ribonucleic acid (RNA) was shed into fecal matter at viral loads high enough to be detected in wastewater [2]. Therefore, the pre-existing field of wastewater-based epidemiology rallied to transition pre-existing methods [3] from academic research into scalable public health surveillance tools. Especially early in the COVID-19 pandemic, traditional swab-based testing could not scale quickly enough to serve as a reliable source of population level disease transmission data [4]. Multiple studies explored the efficacy of WWS as a stream of epidemiological data to complement case tracking for community transmission monitoring, finding that WWS data tracks with trends in clinical case reporting data [5–7]. While the statistical correlation between viral load in sewage and clinical indicators is strong, the exact quantitative relationship between individual-level testing and WWS data is complex and depends on a variety of factors related to the epidemiology of the outbreak as well as data collection and processing timelines [8]. Despite these complexities, WWS has been shown to be correlated with community infection dynamics [7] in addition to simply being an effective qualitative detection tool. Implementing WWS within institutional building complexes, such as college campuses, has unique challenges but also enables building-level resolution monitoring and early warning capabilities [9–12]. WWS can be a leading qualitative indicator of disease presence in a community when overall disease prevalence is low, making WWS a good candidate for broad scale baseline pathogen monitoring [13]. Because WWS is passive and independent of healthcare seeking behavior, it provides a data stream complementary to active tracking of infections or hospitalizations, which both have limitations. Additional benefits of WWS include the ability to monitor multiple pathogens, emerging viral variants, and non-biological hazards [14].

The US Government prioritized WWS to track the spread of COVID-19 and other diseases. For example, environmental monitoring for viral threats via wastewater surveillance is a key component of pandemic threat early-warning systems prioritized in the Biden Administration’s “American Pandemic Preparedness: Transforming our Capabilities” plan [15]. In addition, the Under Secretary of Defense for Personnel and Readiness (USD(P&R)) directed the US Department of Defense (DoD) to leverage alternative technologies, including wastewater surveillance, to supplement existing surveillance strategies in a memorandum titled “Consolidated Department of Defense Coronavirus Disease 2019 Force Health Protection Guidance” [16]. The Centers for Disease Control and Prevention (CDC) established the National Waster Water Surveillance System [17] as a supplement to traditional diagnostic test surveillance systems by enabling efficient collection of community level samples. In addition, CDC is applying WWS within passenger airplanes as part of its Traveler Genomic Surveillance program [18]. Finally, in June 2021, the National Institute of Standards and Technology (NIST) and the Department of Homeland Security (DHS), Science and Technology Directorate convened a virtual workshop, entitled “Standards to Support an Enduring Capability in Wastewater Surveillance for Public Health” to identify challenges and solutions for maturing an ensuring WWS capability for detecting and monitoring public health threats [19,20].

As a result of practical successes in early research and implementation studies, best practices emerged for how to implement WWS at scale [4]. WWS can be an important tool for epidemiological monitoring and outbreak response if implemented with consideration of various challenges [21,22]; one important aspect to consider is avoiding redundancy with clinical testing by implementing a joint surveillance strategy. The design of a WWS data collection scheme and methods for analysis can have significant impacts on bias and interpretation of the data [23]. When implemented according to best practices WWS can be a cost-effective part of a public health response system [24]. Pairing WWS with clinical testing allows for both approaches to serve specific needs thereby enhancing the cost effectiveness of both [25].

The DoD has installations around the globe with small compact living communities, some of which have overlapping watersheds with nearby cities. Tens of thousands of military personnel and civilians live and work in these installations. The DoD implements a four-tiered COVID-19 testing scheme. The first three tiers focus on staff at varying levels of critical service and deployment; Tier 4 sentinel surveillance is an asymptomatic testing program designed to cover all forces. Therefore, we focus on Tier 4 sentinel surveillance as our point of comparison for WWS cost since WWS is also suited to broad population monitoring.

Similarities exist between DoD installations and other institutional building complexes like college campuses. Yet implementing a WWS system at DoD sites requires special planning considerations given unique operational constraints and global scale. To address these issues the DoD commissioned several WWS pilot studies aimed at figuring out the logistical, operational, and financial aspects of implementing a WWS program. One such study demonstrated the effectiveness of wastewater screening of blackwater from Coast Guard vessels [26]. Another study focused on WWS at AFBs; the USAF and JPEO-CBRND WWS pilot study was larger than previous DoD pilots and more representative of US military installations globally. Here we analyze the cost effectiveness of WWS within the DoD context, based on the results from that Air Force and JPEO-CBRND WWS pilot study. We developed a simple cost model that includes upfront capital expenditures, operational expenditures, and indirect costs of lost work time. Further, we perform break-even analysis to explore how traditional swab testing and WWS could be carried out in tandem within the budget of existing swab testing schemes. We conclude that WWS is cost-effective as a complementary passive community level disease surveillance scheme, within the context of AFBs and therefore likely would be cost-effective as a DoD wide global multi-pathogen monitoring system that could be operated in complement to swab-based testing in the event of future disease outbreaks.

## Methods

### WWS pilot study design

To assess feasibility of WWS for SARS-CoV-2 within the USAF context, a multidisciplinary working group was assembled, and a pilot scale implementation was organized. The effort was also coordinated with DoD partners through collaboration with the Office of the Assistant Secretary of Defense for Health Affairs and JPEO-CBRND. A total of 26 AFBs were initially contacted for enrollment either via an invitation from a Public Health Emergency Officer or a USAF Air Staff Logistics Directorate of Civil Engineers memo and all 26 sites expressed initial interest. Wastewater surveillance was ultimately piloted at 17 AFBs to demonstrate feasibility, safety, and utility from May 2021 to January 2022. During the initial phase, conducted June through August 2021, wastewater surveillance techniques were deployed for testing at three remote sites. Next, WWS was evaluated at a larger scale, with 14 additional sites completing standardized procedures to collect and process wastewater samples once per week. The project utilized a portable quantitative polymerase chain reaction (PCR) instrument (qPCR, Biomeme®) [27] and digital PCR (dPCR). AFB site personnel were trained to identify detectable SARS-CoV-2 using both systems. In addition, a passive sampling device was prototyped to decrease costs associated with expensive auto-sampler procurement.

### Collection of tier 4 sentinel surveillance and WWS costs

We gathered known costs or made estimates of direct costs for all activities required to implement both Tier 4 diagnostic testing and WWS protocols. Costs of WWS were based on actual material costs and levels of effort from the WWS pilot study. Costs of Tier 4 diagnostic testing were based directly on USAF experience. Costs included fixed and variable equipment and material costs, and the costs of USAF labor (salaries and benefits) and estimated fully loaded contractor labor (billing) rates. Specific cost parameter values and sources are shown in Table 1.

**Table 1.**
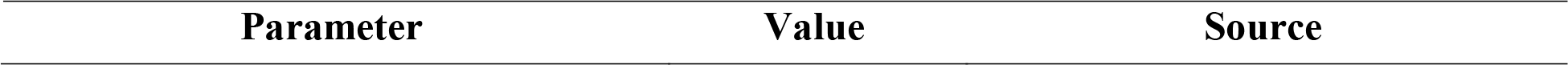
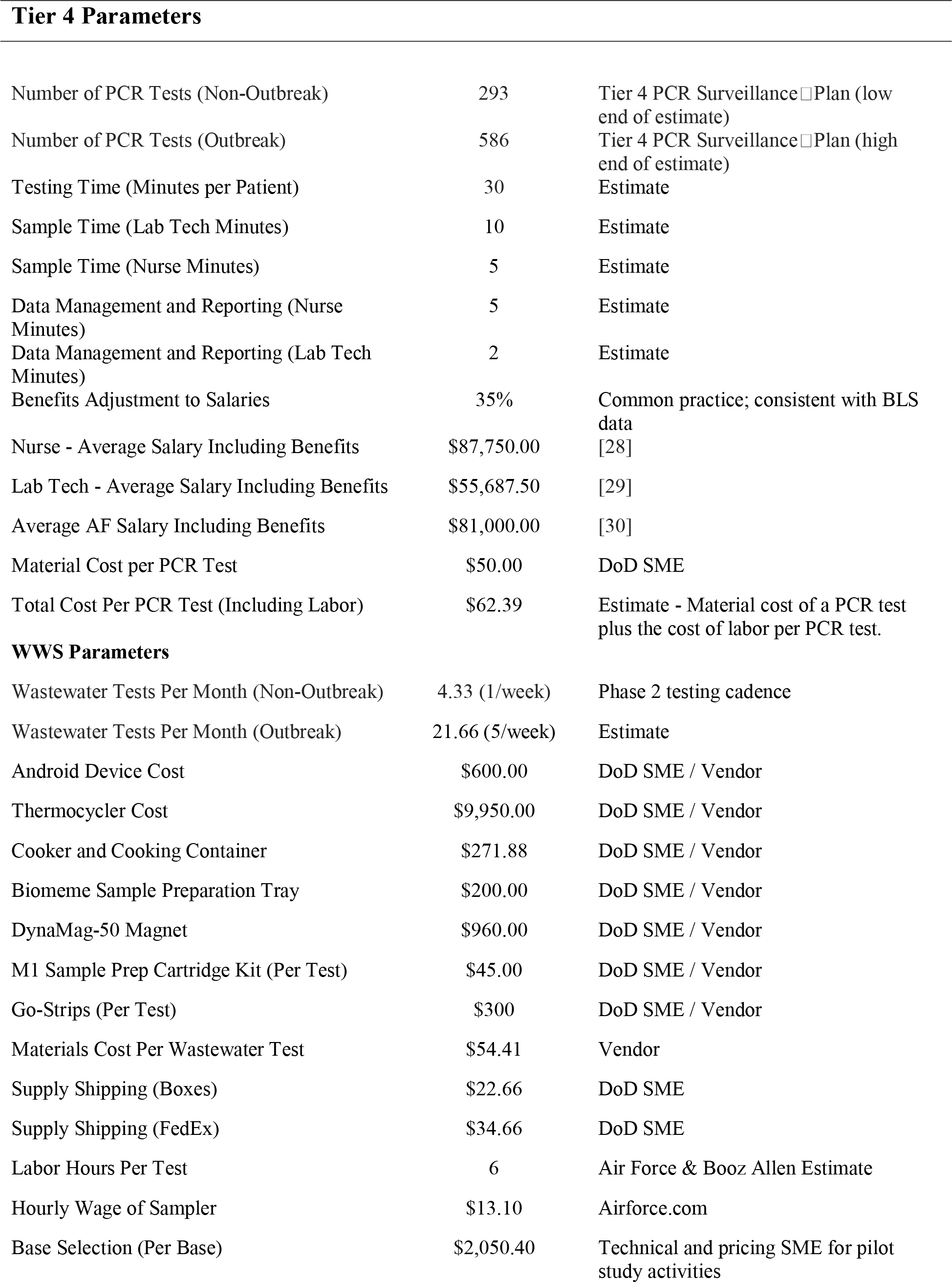
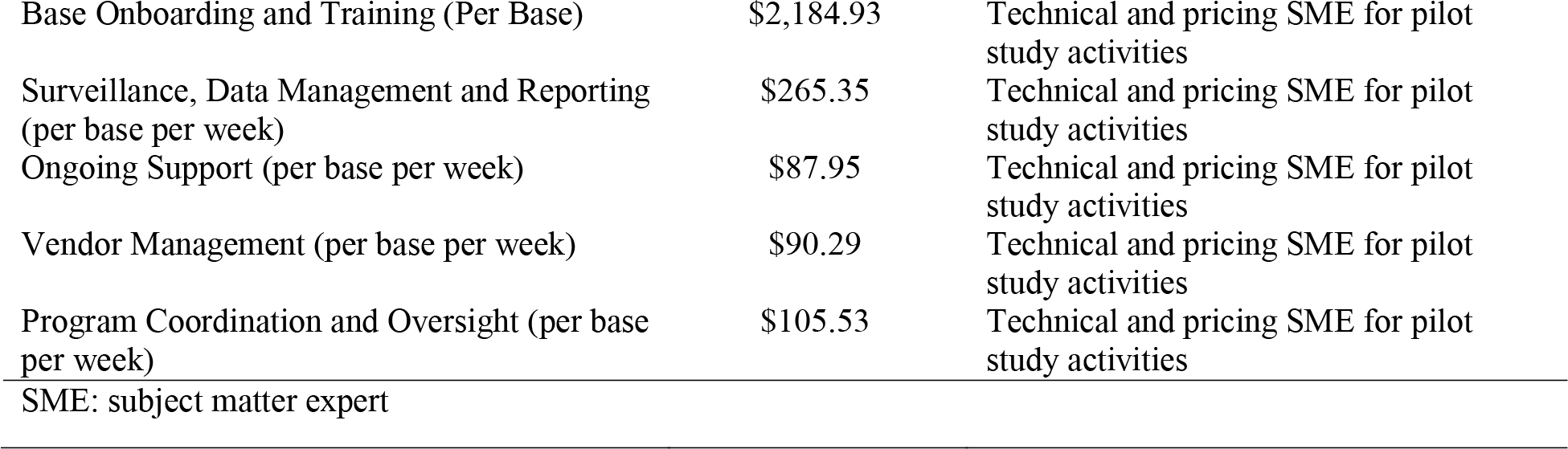
Cost analysis model parameters, with the values used and the sources of the values.

| Parameter | Value | Source |
| --- | --- | --- |
| Tier 4 Parameters |  |  |
| Number of PCR Tests (Non-Outbreak) | 293 | Tier 4 PCR Surveillance □ Plan (low end of estimate) |
| Number of PCR Tests (Outbreak) | 586 | Tier 4 PCR Surveillance □ Plan (high end of estimate) |
| Testing Time (Minutes per Patient) | 30 | Estimate |
| Sample Time (Lab Tech Minutes) | 10 | Estimate |
| Sample Time (Nurse Minutes) | 5 | Estimate |
| Data Management and Reporting (Nurse Minutes) | 5 | Estimate |
| Data Management and Reporting (Lab Tech Minutes) | 2 | Estimate |
| Benefits Adjustment to Salaries | 35% | Common practice; consistent with BLS data |
| Nurse - Average Salary Including Benefits | \$87,750.00 | [28] |
| Lab Tech - Average Salary Including Benefits | \$55,687.50 | [29] |
| Average AF Salary Including Benefits | \$81,000.00 | [30] |
| Material Cost per PCR Test | \$50.00 | DoD SME |
| Total Cost Per PCR Test (Including Labor) | \$62.39 | Estimate - Material cost of a PCR test plus the cost of labor per PCR test. |
| WWS Parameters |  |  |
| Wastewater Tests Per Month (Non-Outbreak) | 4.33 (1/week) | Phase 2 testing cadence |
| Wastewater Tests Per Month (Outbreak) | 21.66 (5/week) | Estimate |
| Android Device Cost | \$600.00 | DoD SME / Vendor |
| Thermocycler Cost | \$9,950.00 | DoD SME / Vendor |
| Cooker and Cooking Container | \$271.88 | DoD SME / Vendor |
| Biomeme Sample Preparation Tray | \$200.00 | DoD SME / Vendor |
| DynaMag-50 Magnet | \$960.00 | DoD SME / Vendor |
| M1 Sample Prep Cartridge Kit (Per Test) | \$45.00 | DoD SME / Vendor |
| Go-Strips (Per Test) | \$300 | DoD SME / Vendor |
| Materials Cost Per Wastewater Test | \$54.41 | Vendor |
| Supply Shipping (Boxes) | \$22.66 | DoD SME |
| Supply Shipping (FedEx) | \$34.66 | DoD SME |
| Labor Hours Per Test | 6 | Air Force & Booz Allen Estimate |
| Hourly Wage of Sampler | \$13.10 | Airforce.com |
| Base Selection (Per Base) | \$2,050.40 | Technical and pricing SME for pilot study activities |
| Base Onboarding and Training (Per Base) | \$2,184.93 | Technical and pricing SME for pilot study activities |
| Surveillance, Data Management and Reporting (per base per week) | \$265.35 | Technical and pricing SME for pilot study activities |
| Ongoing Support (per base per week) | \$87.95 | Technical and pricing SME for pilot study activities |
| Vendor Management (per base per week) | \$90.29 | Technical and pricing SME for pilot study activities |
| Program Coordination and Oversight (per base per week) | \$105.53 | Technical and pricing SME for pilot study activities |
| SME: subject matter expert |  |  |

Materials costs modeled for Tier 4 swab testing only include the total cost of the PCR swab test, which was estimated based on input from DoD subject matter experts with visibility into budgeting and therefore reflects actual cost incurred. The remaining direct Tier 4 costs were associated with labor including the nurse and lab tech time for swab sampling and data management and reporting. We obtained average USAF Nurse base salary values from Salary, a leading industry source of compensation data [28]. Lab tech and USAF general staff salary information were obtained from Indeed [29] and Glassdoor [30] respectively, which are both crowdsourced databases of employers and employees. We assumed a flat benefits rate of 35% and this was added on top of base compensation values to estimate the staff total compensation rates.

WWS labor included a variety of USAF staff and contractors for base and sampling site selection, onboarding bases and training base personnel, obtaining samples, sample processing, data management and reporting, vendor management, and ongoing support to participating bases. The estimate of hourly rate for sample collectors was obtained from publicly available USAF pay tables[31] and contractor rates were estimated based on technical and pricing subject matter expert input and informed by relevant, historical experience.

### Economic cost model

Our analysis addressed the cost of implementing WWS at 82 AFBs, which was the forecasted number of bases expected if a full scale WWS program were to reach maturity [32]. The cost of WWS was evaluated relative to implementing Tier 4 diagnostic testing for the same AFBs. We developed a simple spreadsheet model in Microsoft Excel (Version 2302) to calculate and compare total costs for each surveillance protocol across different scenarios. We assume that there are negligible, if any, startup costs to Tier 4 PCR Surveillance. We also assume that bases would be equipped with suitable resources for Tier 4 surveillance since the pandemic spurred those initial investments. Expected time for staff to go an obtain a clinical PCR test is calculated as the major source of lost work time for Tier 4 surveillance. We assume that there is minimal loss of work under WWS. WWS does not require time spent out of operational environments for staff to get tested, like in Tier 4 diagnostic testing surveillance. Additional staffing required to administer the WWS program and conduct tests is included. The spreadsheet containing the model calculations is provided in **Supplemental Table 1**.

### Cost-effectiveness analysis

We modeled several scenarios to explore the potential costs associated with a range of implementation plans and disease outbreak circumstances. Specifically, the study considered a baseline COVID-19 monitoring scenario (scenario 1), three additional scenarios that explore higher WWS frequency for COVID-19 monitoring (scenario 2), and two scenarios that include increased testing during the 4-month simulated outbreaks (scenarios 3 and 4). For each scenario, the number of outbreak months and number of monthly tests (per base) are described in Table 2. The Air Force Tier 4 sentinel surveillance in practice carried out an average of 293 swab tests per base per month and we assumed that testing would double during outbreaks. The WWS surveillance pilots operated on a once weekly basis, but some evidence exists supporting the benefits of increased sampling frequency. Therefore, we considered increases in baseline sampling and large increases in sampling during outbreaks.

**Table 2.**
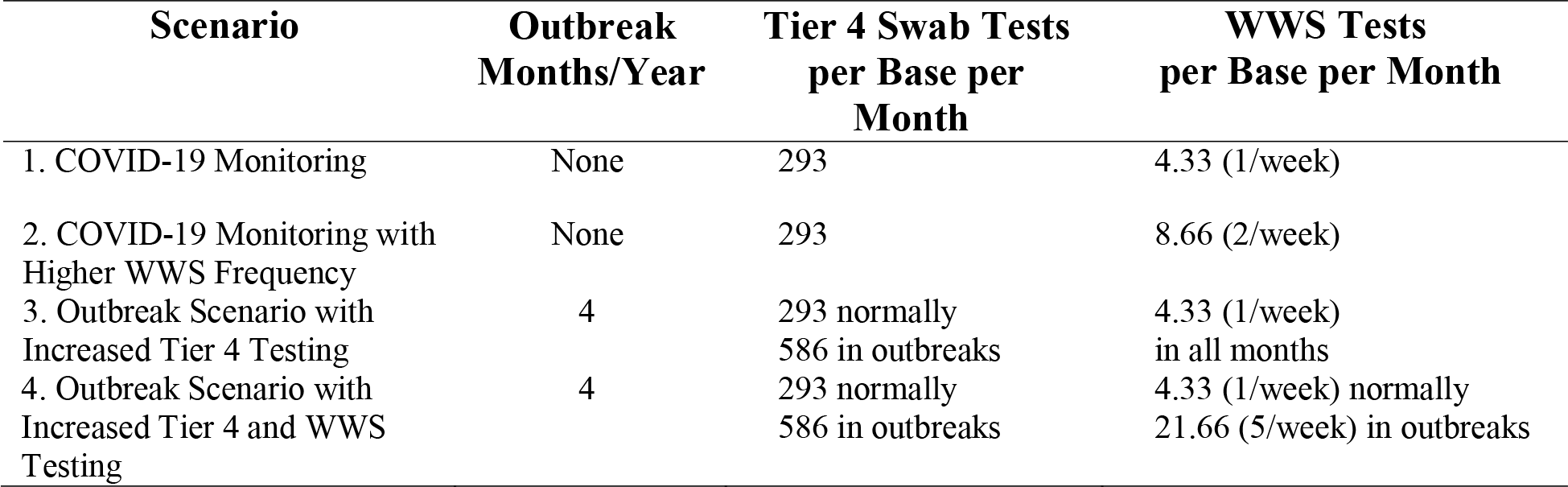
Cost analysis model scenarios.

| Scenario | Outbreak Months/Year | Tier 4 Swab Tests per Base per Month | WWS Tests per Base per Month |
| --- | --- | --- | --- |
| 1. COVID-19 Monitoring | None | 293 | 4.33 (1/week) |
| 2. COVID-19 Monitoring with Higher WWS Frequency | None | 293 | 8.66 (2/week) |
| 3. Outbreak Scenario with Increased Tier 4 Testing | 4 | 293 normally<br>586 in outbreaks | 4.33 (1/week)<br>in all months |
| 4. Outbreak Scenario with Increased Tier 4 and WWS Testing | 4 | 293 normally<br>586 in outbreaks | 4.33 (1/week) normally<br>21.66 (5/week) in outbreaks |

## Results

17 out of the 26 AFBs recorded WWS data during the period from September 2021 to January 2022 (Figure 1); demonstrating that sewage can be safely sampled in a field environment and at a fixed lab. The procedures implemented at sites during the pilot were designed to collect and process wastewater samples once per week. Yet, in total, 52 data submissions were recorded and amounted to 45 unique viable sample records. As illustrated in Figure 2,invalid submissions included duplicate records and samples with PCR reaction issues such incubation temperature and sample concentration. Three sites were used strictly for an early feasibility pilot stage in which protocols were established. The remaining 14 sites submitted data collected over partially overlapping periods of 4.5 weeks on average. In total, the pilot study identified 25 positive (or presumptive positive) samples and 20 negative samples. The 25 positive samples came from 12 of the 14 sites that collected samples systematically. However, the sites did not collect the same number of samples and two sites that detected no positives were also the sites that submitted the fewest total samples. This procedure validated the feasibility of implementing WWS at AFBs, but also highlighted considerable site-to-site variability in executing systematic sampling procedures. These results provided a case study from which we derive assumptions for the economic cost model.

**Figure 1.**
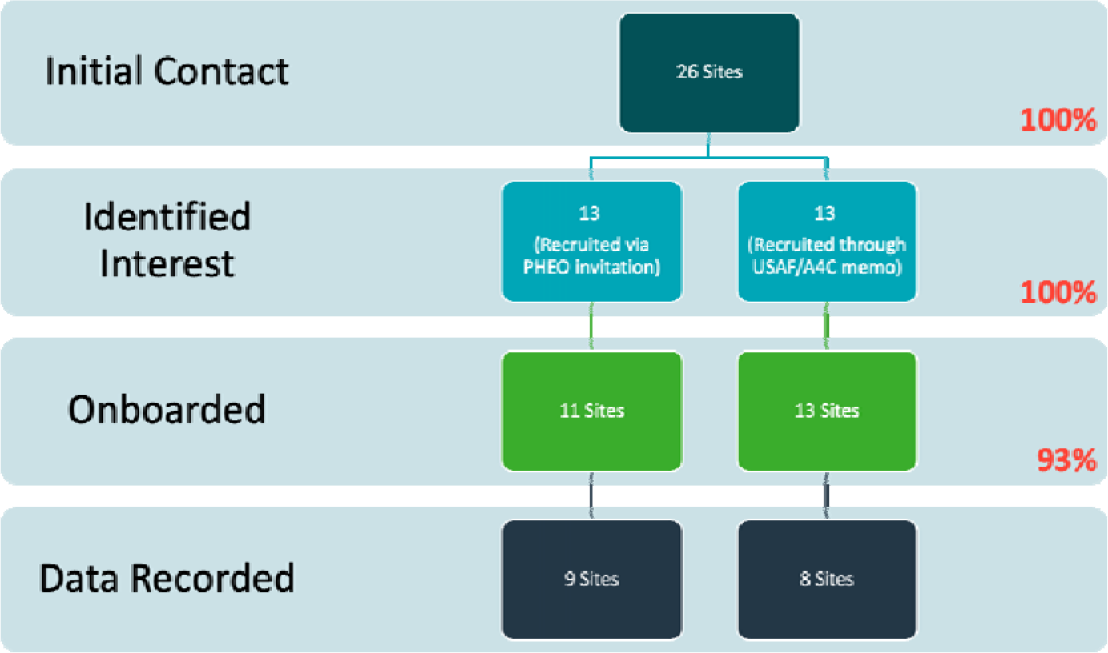
Data collection procedure.

**Figure 2.**
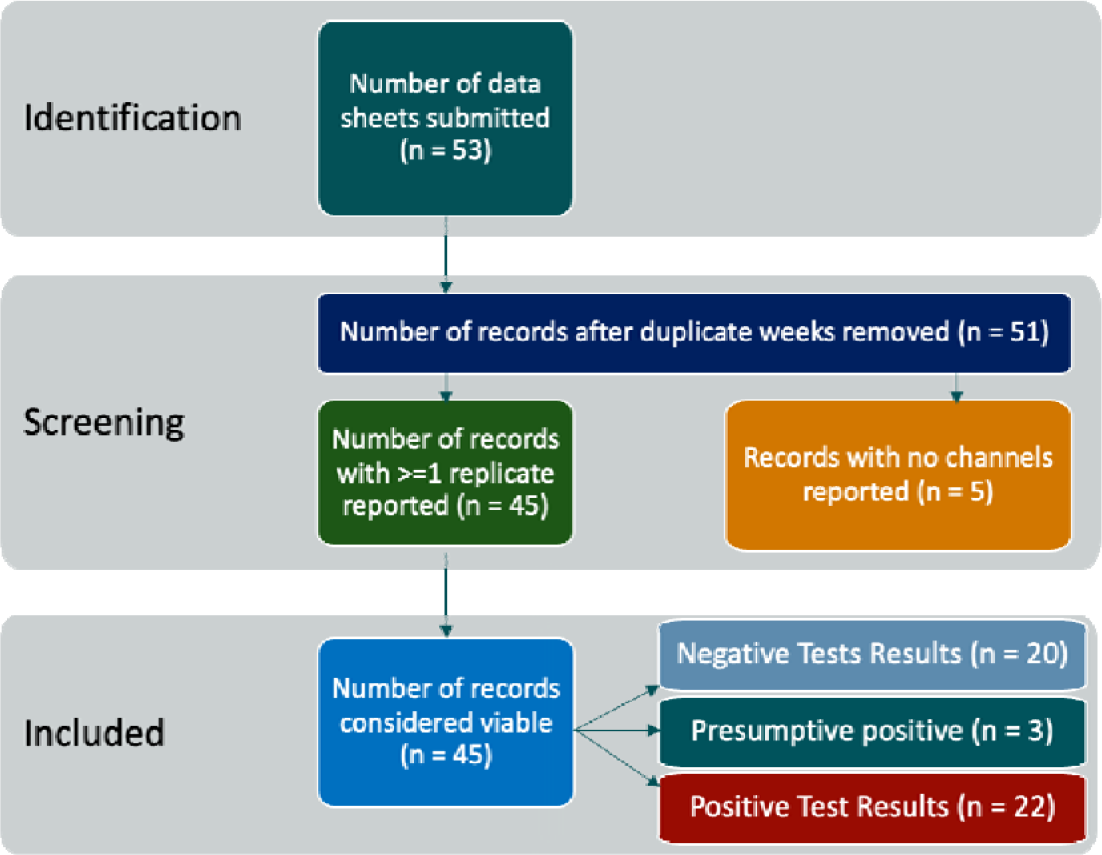
Data preprocessing workflow.

The cost of SARS-CoV-2 WWS was estimated and compared to the estimated cost of Tier 4 COVID-19 sentinel surveillance (asymptomatic testing) across 82 selected AFBs. The four scenarios modeled are described in Table 2. For each scenario we use the cost model parameters to estimate total direct and indirect costs of both WWS and Tier 4 surveillance. Table 3 shows the total annual costs (in millions of 2021 dollars) for each scenario at 82 AFBs. Under baseline COVID-19 monitoring, scenarios 1 and 2, we estimate that the direct costs of the Tier 4 sentinel surveillance program would cost approximately $18 million dollars. In the same scenarios, we estimate that WWS would directly cost between $5.4 million with once weekly testing (scenario 1) to $7.4 million with twice weekly testing (scenario 2).

**Table 3.**
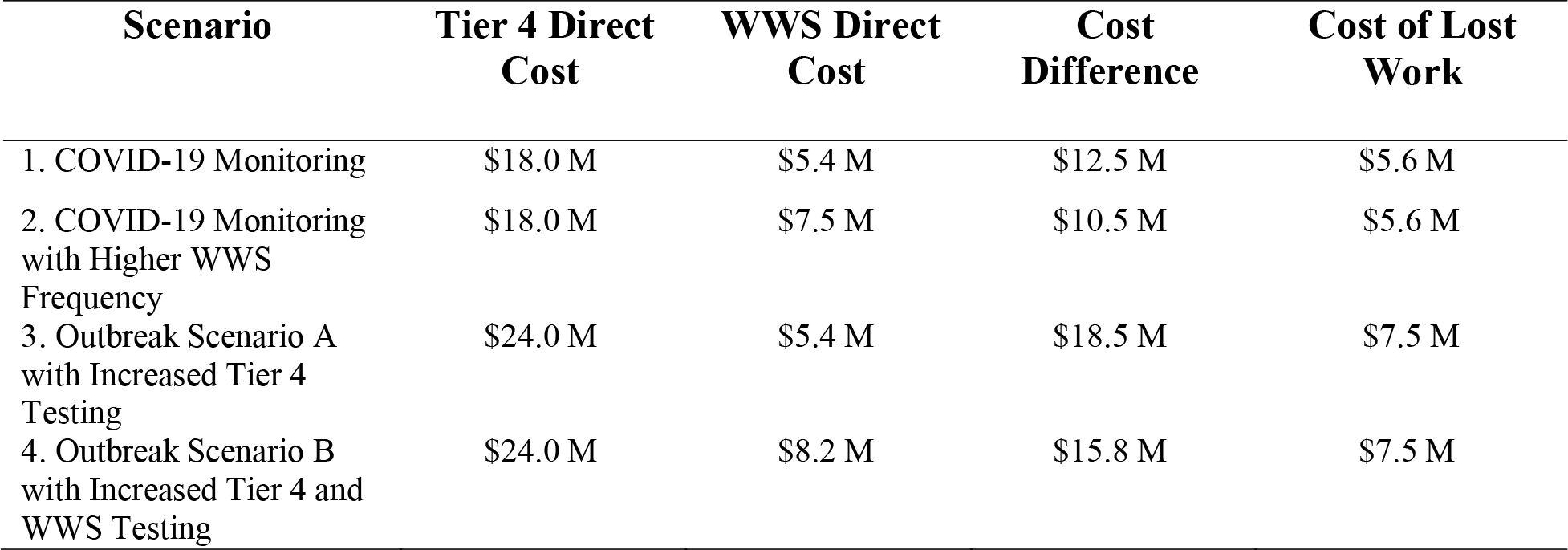
Cost effectiveness analysis results Columns 2 and 3 represent direct costs to administer Tier 4 surveillance and WWS respectively. Column 4 shows the annual cost difference between Tier 4 surveillance and WWS. Column 5 is the indirect cost of staff taking time away from work for Tier 4 surveillance.

During an outbreak, here defined broadly as either local or national level transmission that is sufficiently high such that strict control measures are put in place, enhanced surveillance is needed to manage response. Therefore, under COVID-19 outbreak monitoring scenarios 3 and 4 we estimate the direct costs of Tier 4 sentinel surveillance to be $24 million dollars. The cost of WWS may also go up, depending on policy decision making; for example, sites could choose to test more frequently or utilize alternative pathogen detection methods. In scenario 3, only Tier 4 sentinel surveillance is increased during outbreak response and so the estimated WWS direct costs remain $5.4 million. In contrast, scenario 4 assumed that the usage of both Tier 4 sentinel surveillance and WWS go up during outbreak response and so the estimated WWS direct costs increase to $8.2 million.

Tier 4 sentinel surveillance PCR testing requires that Air Force staff take time out of their workday to get tested and this leads to additional effective costs. We use our model to estimate the cost of lost work, based on typical staff salary ranges and time required to get tested. In scenarios with baseline testing, we estimate that there would be a $5.6 million cost for the loss of work associated with Tier 4 sentinel surveillance PCR testing. During a disease outbreak scenario leading to increased testing, we estimate the cost of lost work to be $7.5 million. In contrast, WWS does not place any burden on staff not associated directly with implementation of the surveillance program.

We find that the cost of WWS was between $10.5 - $18.5 million less expensive annually in direct costs as compared to Tier 4 sentinel surveillance, and that Tier 4 sentinel surveillance has an additional cost of $5.6-$7.5 million annually in Air Force personnel lost work time for testing. If WWS were implemented there would still be capacity to carry out a substantial amount of Tier 4 sentinel surveillance PCR testing. We quantified the break-even point for combined WWS and PCR testing by calculating the number of PCR swab tests that could be conducted per base per month under the WWS paradigm while breaking even with the higher cost of the original Tier 4 testing scheme. The results of our break-even analysis are shown in Table 4.

**Table 4.** Breakeven analysis results.

| Scenario | Breakeven Swab Tests | Breakeven Swab Tests |
| --- | --- | --- |

|  | **Based on Direct Cost**<br>**Difference Only<sup>1</sup>**<br>**(per AFB/month)** | **Including Cost of Lost**<br>**Work<sup>1</sup>**<br>**(per AFB/month)** |
| --- | --- | --- |
| 1. COVID-19 Monitoring | 204 | 225 |
| 2. COVID-19 Monitoring with Higher<br>WWS Frequency | 171 | 200 |
| 3. Outbreak Scenario with Increased<br>Tier 4 Testing | 302 | 323 |
| 4. Outbreak Scenario with Increased<br>Tier 4 and WWS Testing | 258 | 289 |

<sup>1</sup>Swab tests per base per month added to WWS at the breakeven point relative to Tier 4 costs; in outbreak scenarios, these figures reflect a full year (4 outbreak months and 8 non-outbreak months).

To estimate break-even point based on direct costs only, we simply take the direct cost differences and divide by the direct cost per swab test ($62.39/test for materials and labor), spread across 82 bases and 12 months. Under COVID-19 baseline monitoring with once weekly WWS (scenario 1) we estimate that an additional 204 swab tests per AFB per month could be added to the WWS protocol for the same cost as the original Tier 4 sentinel surveillance scheme. If WWS was increased to twice weekly (scenario 2) then we estimate that 171 swab tests could still be performed at the break-even point. When cost of lost work is incorporated, including assumed lost work for swabs used to reach break-even point, we estimate that 225 and 200 additional swab tests could be performed in scenarios 1 and 2 respectively. During an outbreak, the demands for all forms of surveillance increases and so more swab tests can be performed at the break-even cost point. When the cost of lost work is included, we estimate that 323 and 289 additional swab tests could be performed for outbreak scenarios 3 and 4 respectively. Across all scenarios, we estimate that more than half of the Tier 4 sentinel surveillance program could be maintained while WWS is implemented in parallel with no additional cost, i.e., at the break-even point.

## Discussion

The DoD SARS-CoV-2 WWS surveillance pilot studies demonstrated the feasibility of implementing WWS at military installations. The pilot studies revealed some important technical considerations. For example, although dPCR was extremely sensitive, it required shipping of wastewater from remote sites to a centralized location, potentially limiting its use in largescale deployment. Portable qPCR had a lower throughput of samples than dPCR but was simple to use at the point of sampling. Our simple cost model took these lessons into account.

The pilot studies also provide real world data on the costs associated with WWS in comparison to standard swab-based testing, including materials costs and labor requirements. In general, WWS for SARS-CoV-2 may offer several benefits, including 1) earlier detection of outbreaks, 2) lower cost and burden for community-wide coverage compared to diagnostic testing, and 3) detection of viral presence, regardless of symptoms. Coupling our analysis with the overall results of the DoD pilot studies suggests that those benefits are likely to transfer to the DoD. Specifically, our model suggests deployment of WWS to AFBs would be substantially more cost effective than broad asymptomatic swab testing. Our break-even analysis indicates that without allocating additional funding to surveillance efforts that WWS could be implemented for AFB level monitoring and swab testing could be used for more targeted purposes or simply in parallel. It is important to note that swab testing and WWS do not provide the exact same information. Swab testing can enable individual level actions, such as quarantining and contract tracing, and higher resolution data. Therefore, tradeoffs between the public health benefits of WWS and swab testing will need to be determined on a case-by-case basis. These findings apply do both baseline COVID-19 monitoring and in scenarios where outbreaks are occurring on bases throughout the year.

Our cost model was intentionally simplistic to enhance transparency for decisionmakers. That simplicity is also a limitation in that there may be unforeseen complexities and costs associated with scaling the WWS program beyond the pilot sites. In addition, our cost data is primarily derived from the Air Force WWS pilot program, and it is possible that facilities associated with other branches of the DoD may require different considerations. Many of our parameter estimates were obtained from subject matter experts, i.e., individual DoD staff and contractors associated with the pilot studies, as opposed to an independent review of pilot study budgets. Therefore, our cost estimates should not be interpreted as formal financial forecasts.

Another limitation of our analysis is the lack of uncertainty estimation. Any formal program level financial forecast would require uncertainty ranges to be estimated along with point cost estimates. However, in our present work many of the materials costs were obtained directly from individuals with knowledge of the actual costs incurred during the pilot studies. Therefore, our model could be framed as an estimate of what the actual cost would have been if the pilot was carried out at all AFBs rather than a forecast of the costs of a DoD wide program—although we believe our work is germane to that topic. Furthermore, systematic uncertainty in labor and materials costs due to changes in supply chain issues and inflation are likely correlated such that a proper uncertainty propagation would require estimating the joint distribution of costs, which is beyond the scope of our efforts. Given the magnitude of the point difference and the consensus in the literature that WWS is less expensive for population level monitoring—albeit not necessarily cost-effective if implemented poorly [22]--we believe that our results are qualitatively robust to underlying uncertainty in the data and model specification.

In conclusion, we find that the Air Force WWS pilot was a cost-effective complement to standard swab-based testing as implemented in the Tier 4 sentinel surveillance program. We believe that WWS and swab-based testing have differing strengths and weaknesses; implementing both approaches in tandem offers the opportunity to specialize. Looking ahead beyond the COVID-19 pandemic the DoD can be an important partner in global pandemic and all-hazard preparedness efforts. WWS is uniquely well suited to multi-threat biological surveillance and our results suggest that adoption of WWS across US military installations would help deliver a more comprehensive early warning system. A DoD wide WWS would complement civilian efforts like the National Wastewater Surveillance System and enable rapidly scalable outbreak monitoring in the event of future disease outbreaks.

## Data Availability

All data produced in the present work are contained in the manuscript

## Acknowledgements

We would like to thank Helen Phipps for helpful input. JDV and SS designed the study. EMM managed the study and carried out the analysis. RP, JT, and EJM carried out the analysis. JS wrote the manuscript.

## Disclaimer

The views expressed in this manuscript are those of the authors and do not necessarily reflect the official policy or position of the Air Force, the Joint Program Executive Office for Chemical, Biological, Radiological and Nuclear Defense (JPEO-CBRND), the Department of Defense, or the U.S. Government. Included references to commercial products do not constitute endorsement by the Department of Defense.

## References

1. Sinclair RG, Choi CY, Riley MR, Gerba CP. Pathogen surveillance through monitoring of sewer systems. Adv Appl Microbiol 2008;65:249–269. PMID:19026868

2. Medema G, Heijnen L, Elsinga G, Italiaander R, Brouwer A. Presence of SARS-Coronavirus-2 RNA in Sewage and Correlation with Reported COVID-19 Prevalence in the Early Stage of the Epidemic in The Netherlands. Environ Sci Technol Lett 2020 May 20;acs.estlett.0c00357. PMID:null

3. Sims N, Kasprzyk-Hordern B. Future perspectives of wastewater-based epidemiology: Monitoring infectious disease spread and resistance to the community level. Environment International 2020 Jun 1;139:105689. doi: 10.1016/j.envint.2020.105689

4. Keshaviah A, Hu XC, Henry M. Developing a Flexible National Wastewater Surveillance System for COVID-19 and Beyond. Environmental Health Perspectives 2021 Apr 20;129(4):045002. doi: 10.1289/EHP8572

5. Ahmed W, Tscharke B, Bertsch PM, Bibby K, Bivins A, Choi P, Clarke L, Dwyer J, Edson J, Nguyen TMH, O’Brien JW, Simpson SL, Sherman P, Thomas KV, Verhagen R, Zaugg J, Mueller JF. SARS-CoV-2 RNA monitoring in wastewater as a potential early warning system for COVID-19 transmission in the community: A temporal case study. Sci Total Environ 2021 Mar 20;761:144216. PMID:33360129

6. Feng S, Roguet A, McClary-Gutierrez JS, Newton RJ, Kloczko N, Meiman JG, McLellan SL. Evaluation of Sampling, Analysis, and Normalization Methods for SARS-CoV-2 Concentrations in Wastewater to Assess COVID-19 Burdens in Wisconsin Communities. ACS EST Water 2021 Aug 13;1(8):1955–1965. doi: 10.1021/acsestwater.1c00160

7. Peccia J, Zulli A, Brackney DE, Grubaugh ND, Kaplan EH, Casanovas-Massana A, Ko AI, Malik AA, Wang D, Wang M, Warren JL, Weinberger DM, Arnold W, Omer SB. Measurement of SARS-CoV-2 RNA in wastewater tracks community infection dynamics. Nat Biotechnol Nature Publishing Group; 2020 Oct;38(10):1164–1167. doi: 10.1038/s41587-020-0684-z

8. Olesen SW, Imakaev M, Duvallet C. Making waves: Defining the lead time of wastewater-based epidemiology for COVID-19. Water Res 2021 Sep 1;202:117433. PMID:34304074

9. Betancourt WQ, Schmitz BW, Innes GK, Prasek SM, Pogreba Brown KM, Stark ER, Foster AR, Sprissler RS, Harris DT, Sherchan SP, Gerba CP, Pepper IL. COVID-19 containment on a college campus via wastewater-based epidemiology, targeted clinical testing and an intervention. Sci Total Environ 2021 Jul 20;779:146408. PMID:33743467

10. Karthikeyan S, Nguyen A, McDonald D, Zong Y, Ronquillo N, Ren J, Zou J, Farmer S, Humphrey G, Henderson D, Javidi T, Messer K, Anderson C, Schooley R, Martin NK, Knight R. Rapid, Large-Scale Wastewater Surveillance and Automated Reporting System Enable Early Detection of Nearly 85% of COVID-19 Cases on a University Campus. mSystems American Society for Microbiology; 2021 Aug 10;6(4):e00793–21. doi: 10.1128/mSystems.00793-21

11. Landstrom M, Braun E, Larson E, Miller M, Holm GH. Efficacy of SARS-CoV-2 wastewater surveillance for detection of COVID-19 at a residential private college. FEMS Microbes 2022 Jan 1;3:xtac008. doi: 10.1093/femsmc/xtac008

12. Scott LC, Aubee A, Babahaji L, Vigil K, Tims S, Aw TG. Targeted wastewater surveillance of SARS-CoV-2 on a university campus for COVID-19 outbreak detection and mitigation. Environ Res 2021 Sep;200:111374. PMID:34058182

13. Kirby AE. Using Wastewater Surveillance Data to Support the COVID-19 Response — United States, 2020–2021. MMWR Morb Mortal Wkly Rep 2021;70. doi: 10.15585/mmwr.mm7036a2

14. Li L, Mazurowski L, Dewan A, Carine M, Haak L, Guarin TC, Dastjerdi NG, Gerrity D, Mentzer C, Pagilla KR. Longitudinal monitoring of SARS-CoV-2 in wastewater using viral genetic markers and the estimation of unconfirmed COVID-19 cases. Sci Total Environ 2022 Apr 15;817:152958. PMID:35016937

15. Lander ES, Sullivan JJ. American Pandemic Preparedness: Transforming Our Capabilities. Available from: whitehouse.gov

16. CONSOLIDATED-DEPARTMENT-OF-DEFENSE-CORONAVIRUS-DISEASE-2019-FORCE-HEALTH-PROTECTION-GUIDANCE-REVISION-5.pdf. Available from: https://media.defense.gov/2023/Mar/28/2003187831/-1/-1/1/CONSOLIDATED-DEPARTMENT-OF-DEFENSE-CORONAVIRUS-DISEASE-2019-FORCE-HEALTH-PROTECTION-GUIDANCE-REVISION-5.PDF [accessed May 24, 2023]

17. National Wastewater Surveillance System. Centers for Disease Control and Prevention. 2023. Available from: https://www.cdc.gov/nwss/wastewater-surveillance.html [accessed May 4, 2023]

18. Traveler-Based Genomic Surveillance for Early Detection of New SARS-CoV-2 Variants | Travelers’ Health | CDC. Available from: https://www.nc.cdc.gov/travel/page/travel-genomic-surveillance [accessed Sep 25, 2023]

19. Lin N, Servetas S, Jackson S, Lippa K, Parratt K, Mattson P, Beahn C, Mattioli M, Gutierrez S, Focazio M, Smith T, Storella P, Wright S. Report on the DHS/NIST Workshop on Standards for an Enduring Capability in Wastewater Surveillance for Public Health (SWWS Workshop). NIST Nancy Lin, Stephanie Servetas, Scott Jackson, Katrice Lippa, Kirsten Parratt, Philip Mattson, Clare Beahn, Mia Mattioli, Sally Gutierrez, Michael Focazio, Ted Smith, Paul Storella, Sarah Wright; 2022 Aug 16; Available from: https://www.nist.gov/publications/report-dhsnist-workshop-standards-enduring-capability-wastewater-surveillance-public [accessed May 24, 2023]

20. Servetas SL, Parratt KH, Brinkman NE, Shanks OC, Smith T, Mattson PJ, Lin NJ. Standards to support an enduring capability in wastewater surveillance for public health: Where are we? Case Studies in Chemical and Environmental Engineering 2022 Dec;6:100247. PMID:null

21. O’Keeffe J. Wastewater-based epidemiology: current uses and future opportunities as a public health surveillance tool. Environ Health Rev 2021 Nov;64(3):44–52. doi: 10.5864/d2021-015

22. Safford HR, Shapiro K, Bischel HN. Wastewater analysis can be a powerful public health tool—if it’s done sensibly. Proceedings of the National Academy of Sciences 2022 Feb 8;119(6):e2119600119. doi: 10.1073/pnas.2119600119

23. Safford H, Zuniga-Montanez RE, Kim M, Wu X, Wei L, Sharpnack J, Shapiro K, Bischel HN. Wastewater-Based Epidemiology for COVID-19: Handling qPCR Nondetects and Comparing Spatially Granular Wastewater and Clinical Data Trends. ACS EST Water 2022 Nov 11;2(11):2114–2124. doi: 10.1021/acsestwater.2c00053

24. Hart OE, Halden RU. Computational analysis of SARS-CoV-2/COVID-19 surveillance by wastewater-based epidemiology locally and globally: Feasibility, economy, opportunities and challenges. Science of The Total Environment 2020 Aug 15;730:138875. doi: 10.1016/j.scitotenv.2020.138875

25. Wright J, Driver EM, Bowes DA, Johnston B, Halden RU. Comparison of high-frequency in-pipe SARS-CoV-2 wastewater-based surveillance to concurrent COVID-19 random clinical testing on a public U.S. university campus. Sci Total Environ 2022 May 10;820:152877. PMID:34998780

26. Hall GJ, Page EJ, Rhee M, Hay C, Krause A, Langenbacher E, Ruth A, Grenier S, Duran AP, Kamara I, Iskander JK, Thomas DL, Bock E, Porta N, Pharo J, Osterink BA, Zelmanowitz S, Fleischmann CM, Liyanage D, Gray JP. Wastewater Surveillance of U.S. Coast Guard Installations and Seagoing Military Vessels to Mitigate the Risk of COVID-19 Outbreaks. medRxiv; 2022. p. 2022.02.05.22269021. doi: 10.1101/2022.02.05.22269021

27. Biomeme SARS-CoV-2 Real-Time RT-PCR Test - Letter of Authorization | FDA. Available from: https://www.fda.gov/media/141049 [accessed May 24, 2023]

28. Salary.com S built by: U.S. Air Force (USAF) Nurse Salary. Salary.com. Available from: https://www.salary.com/research/salary/employer/u-s-air-force-usaf/nurse-salary [accessed Oct 19, 2023]

29. Laboratory technician salary in United States. Available from: https://www.indeed.com/career/laboratory-technician/salaries [accessed Oct 19, 2023]

30. How Much Does US Air Force Pay in 2023? (41,869 Salaries). Glassdoor. Available from: https://www.glassdoor.com/Salary/US-Air-Force-Salaries-E41283.htm [accessed Oct 19, 2023]

31. Air Force Active Duty Benefits - U.S. Air Force. Available from: https://www.airforce.com/pay-and-benefits/air-force-benefits [accessed Oct 19, 2023]

32. Locations - U.S. Air Force. Available from: https://www.airforce.com/ways-to-serve/locations [accessed May 24, 2023]

